# Temporal Co-Evolution of Clinical Scores and Neuro-Immune Biomarkers in Preterm Infants with Severe Germinal Matrix-Intraventricular Hemorrhage and Post-Hemorrhagic Ventricular Dilation

**DOI:** 10.64898/2026.06.02.26349284

**Authors:** Sara Bitarafan, Angel del Marco, Isabel Benavente-Fernandez, Juan Arnaez, Simon Lubian-Lopez, Levi B. Wood, Monica Garcia-Alloza

## Abstract

Germinal matrix-intraventricular hemorrhage (GM-IVH) is one of the most frequent and severe neurological complications in preterm infants (PT). It triggers an inflammatory response accompanied by neuronal and glial injury and may progress to post-hemorrhagic ventricular dilatation (PHVD), thereby increasing long term disability and cognitive deficits. Nevertheless, the characteristics and evolution of the associated pathology is poorly understood. To assess neuroimmune response and neuropathology induced by GM-IVH, we quantified cytokines, glial activation and neurodegeneration makers in cerebrospinal fluid collected from 12 patients with grades III/IV GM-IVH and PHVD and 5 controls neonates from the onset of pathology up to 2 months of age. Additionally, to evaluate long-term deficits and behavioral outcomes, we used standard behavioral test including Bayley Scales of Infant and Toddler Development, Third Edition (Bayley-III) at 2 years of age. Interestingly, we found that while pathology markers such as ubiquitin carboxy-terminal hydrolase L1 (UCHL1), alpha II spectrin breakdown product 145 (SBDP145) and myelin basic protein (MBP) are elevated in PT, their level decline over time. Furthermore, cytokine profiling identified two divergent temporal trajectories (i.e., diminishing or sustained) that correspond with either neuronal or astrocytic markers. Specifically, diminishing cytokines including IL-6, IL-8, and IP-10 decreased with age and were correlated with neuronal markers such as SBDP145, UCH-L1, and MBP. In contrast, sustained cytokines such as IFN-γ, IL-7, IL-13, and MCP-1 remained elevated or unchanged throughout the study period and were positively correlated with astrocyte reactivity marker GFAP. Notably, sustained cytokines were consistent with worse motor function and behavioral outcome. Together, longitudinal CSF analysis in PT with severe GM-IVH and PHVD identifies a cytokine profile that declines and correlates with neuronal and glial injury markers, and another that remains sustained and correlates with gliosis and adverse neurodevelopmental outcomes. These findings highlight potential CSF biomarkers associated with disease progression and long-term neurological impairment, providing a foundation for future evaluation of candidate therapeutic interventions.

## INTRODUCTION

Complications related to preterm birth (<37 gestational weeks) are the leading cause of death among children under five years of age, accounting for approximately one million deaths in 2015 (WHO, 2018). According to the World Health Organization, approximately 15 million babies are born prematurely each year worldwide (WHO, 2018), accounting for ≈11% of all births (Vogel et al., 2018). Although the prevalence of preterm birth varies substantially across countries and regions (Blencowe et al., 2019; Chawanpaiboon et al., 2019; Walani, 2020), its overall rate has been increasing in recent decades Importantly, prematurity is associated with many short- and long-term complications in surviving preterm infants (PT) (Behrman, Butler, & Institute of Medicine (U.S.). Committee on Understanding Premature Birth and Assuring Healthy Outcomes., 2007; Cassiano, Gaspardo, & Linhares, 2016; Pravia & Benny, 2020). Among the various life-threatening complications, germinal matrix–intraventricular hemorrhage (GM-IVH) is one of the most severe conditions affecting PT (Ballabh, 2010; Chevallier et al., 2017), typically occurring within the first 48 hours after birth (Ballabh, 2010; Kuo, 2020). GM-IVH is caused by rupture and subsequent hemorrhage of the immature and fragile blood vessels of the germinal matrix that may disrupt the ependymal tissue layer and extend to the ventricles (Egesa et al., 2021; Smith, 2008; Volpe, 2015). Additionally, severe GM-IVH (grades III/IV) occurs in a substantial proportion of infants that may reach up to one half of PT in some studies, involving bleeding into the ventricular system, which leads to post-hemorrhagic ventricular dilatation (PHVD) (El-Dib et al., 2020; Kuo, 2020).

GM-IVH may result in short and long-term complications, including cerebral palsy, motor impairment, or cognitive deficits (Atienza-Navarro, Alves-Martinez, Lubian-Lopez, & Garcia-Alloza, 2020; Hee Chung, Chou, & Brown, 2020), which are reported in 45-86% of PT with severe GM-IVH (McCrea & Ment, 2008), and are accentuated by progressive PHVD (Adams-Chapman, Hansen, Stoll, Higgins, & Network, 2008; Dorner et al., 2019; Srinivasakumar et al., 2013; Volpe, 2020). Although the precise causes of progressive PHVD following GM-IVH remain largely unclear, ventriculomegaly and associated brain tissue damage observed in these patients may primarily result from cytokine-driven inflammatory responses which can lead to abnormal cerebrospinal fluid (CSF) drainage and exacerbate periventricular injury, underscoring cytokine profile as both a key pathophysiological mechanism and a promising therapeutic target (Gilard, Tebani, Bekri, & Marret, 2020; Holste, Xia, Ye, Keep, & Xi, 2022; Karimy et al., 2017; J. Li, Zhang, Guo, Yu, & Yang, 2021; Limbrick et al., 2017; Magid-Bernstein et al., 2025). Besides, CSF pressure can rise to approximately 9.1 mmHg in PT with PHVD, in contrast to a typical range of 2.8-5.7 mmHg in healthy neonates (Kaiser & Whitelaw, 1985). In severe cases, a temporary ventricular reservoir or shunt must be placed to limit CSF accumulation and the consequent increase in intracranial pressure and compression of the brain parenchyma (de Vries et al., 2002; El-Dib et al., 2020; Gaskill, Marlin, & Rivera, 1988; Mahoney, Luyt, Harding, & Odd, 2020; Murphy et al., 2002; J. Y. Wang, Amin, Jallo, & Ahn, 2014). While these provide symptomatic relief, GM-IVH has no successful treatment to limit brain damage and subsequent neurodevelopmental complications.

Additionally, rupture of the blood–brain barrier (BBB) in the germinal matrix and the subsequent hemorrhage trigger an inflammatory response and allow the entry of toxic blood-derived factors, including erythrocytes that release their cytoplasmic contents (Chen et al., 2015; B. Cheng & Ballabh, 2022; Gram et al., 2013; Holste et al., 2022; Strahle et al., 2021). This cascade leads to increased levels of reactive oxygen species, pro-inflammatory cytokines, white blood cells, macrophages, and activation of microglia, all of which accumulate in the CSF within the ventricular space. (Chari, Mallucci, Whitelaw, & Aquilina, 2021; Chen et al., 2015; B. Cheng & Ballabh, 2022; Durocher et al., 2021; Holste et al., 2022; Otun et al., 2021). However, as far as we know, no previous study has longitudinally monitored neuronal and inflammatory markers in the CSF of PT with GM-IVH and PHVD Given the potential for brain inflammation to drive tissue degeneration, in the current study, we sought to determine if the immune milieu in the CSF of PT patients who suffered from GM-IVH and PHVD is related to pathological outcomes. We analyzed CSF from PT with GM-IVH and PHVD and quantified a profile of inflammatory cytokines and markers of neuronal and glial damage with time from birth up to 2 months of life. We correlated these markers with anatomical and cognitive parameters. Importantly, we have found that certain inflammatory markers are acutely increased, such as interferon-γ-inducible protein (IP)-10 and interleukin (IL)-8 in PT, while the levels of other cytokines, such as monocyte chemoattractant protein (MCP)-1 and IL-17, remain constant with time. Moreover, these changes also relate to neuron damage, as analyzed by neurofilament light chain (NF-light) or phosphorylated-tau levels. Overall, to our knowledge this study characterizes for the first time immune and pathological markers in PT and evaluates how temporal inflammatory changes relate to long-term outcomes. This study provides foundation for future non-invasive therapeutic interventions.

## MATERIAL AND METHODS

### Research subjects and study design

We retrospectively analyzed CSF from preterm infants <32 weeks of post-menstrual age with progressive PHVD and preterm infants who underwent lumbar puncture for sepsis evaluation to rule out meningitis. Samples were collected at Puerta del Mar University Hospital between February 2013 and April 2020. PHVD was established by repeated cerebral ultrasonography when GM-IVH was followed by a progressive increase in ventricular width above 97th centile. Inclusion criteria: (1) preterm infants <32 weeks estimated post-menstrual age with birth weight *<*1500 grams and a grade III or IV GM-IVH diagnosed with cerebral ultrasonography; (2) <28 days after birth; (3) progressive dilatation of both lateral ventricles with ventricular width >p97 and anterior horn width >6 mm; (4) ventricular reservoir (VR) insertion. Exclusion criteria: (1) chromosomal disorders; (2) congenital malformations; (3) cystic periventricular leukomalacia; (4) infection of the central nervous system; (5) primary caregiver refusing participation. PT with no known neurologic disease who underwent a lumbar puncture for sepsis evaluation, where the evaluation and all cultures were negative, were included as controls. **Table 1** shows the demographic and clinical data of the patients included in the study. All collected data are in **Table S1.**

**Table 1.** Patient Demographic and Clinical Characteristics.

| Demographic and clinical characteristics of patients | PT<br>(n=8-12) | Control<br>(n=5) |
| --- | --- | --- |
| Grade III/IV GM-IVH and PHVD, No. (%) | 12 (100) | 0 (0) |
| Birth weight, mean (SEM), g | 1158,9 (91,7) | 3379,8<br>(368,9) |
| Gestational age at birth, median (IQR), week | 27 (3,5) | 39 (2,5) |
| Sex, No. (%) |  |  |
| Female | 6 (50) | 3 (60) |
| Male | 3 (25) | 2 (40) |
| Undetermined | 3 (25) | - |
| Multiple gestation (twins), No. (%) | 2 (22,2) | 0 (0) |
| Vaginal delivery, No. (%) | 7 (77,7) | 2 (40) |
| BPD, No. (%) | 4 (50) | 0 (0) |
| Apgar score at 1 min, median (IQR) | 6 (1,5) | 9 (1) |
| Apgar score at 5 min, median (IQR) | 8 (1) | 10 (1) |
| CRIB score, median (IQR)* | 3 (4) | - |
| Ventricle peritoneal shunt, No. (%) | 10 (83,3) | 0 (0) |
| Death | 2 (18,2) | 0 (0) |
| Head circumference, mean (±SEM), cm | 27,1 (0,8) | 35,6 (0,6) |
| Ultrasound measurements*<br>(First measure, when the pathology appears) |  |  |
| Right VV, mean (±SEM), cc (n=7) | 12,8 (4,1) | - |
| Left VV, mean (±SEM), cc (n=6) | 15,6 (5,3) | - |
| TBV, mean (±SEM), cc (n=7) | 198,5 (22,5) | - |
| IVD, mean (±SEM), mm (n=11) | 14,4 (1,1) | - |
| IVI, mean (±SEM), mm (n=10) | 14,8 (1,3) | - |
| AHWD, mean (±SEM), mm (n=11) | 8,75 (1,6) | - |
| AHWI, mean (±SEM), mm (n=10) | 10,7 (1,8) | - |
| TODD, mean (±SEM), mm (n=10) | 23,9 (1,9) | - |
| TODI, mean (±SEM), mm (n=9) | 25,9 (2,1) | - |
| Bayley Scales of Infant Development and prognosis at 2 years of age* |  |  |
| Cognition, mean (±SEM) (n=6) | 90 (4) | - |
| Cognition score <70, No. (%) | 0 (0) | - |
| Language, mean (±SEM) (n=6) | 93 (5,6) | - |
| Language score <70, No. (%) | 0 (0) | - |
| Motor assessment, mean (±SEM) (n=6) | 41 (18,9) | - |
| Motor assessment score <70, No. (%) | 4 (66,7) | - |
| Societal-emotional assessment, mean (±SEM) (n=6) | 35,5 (19,3) | - |
| Societal-emotional assessment score <70, No. (%) | 4 (66,7) | - |
| Adaptation assessment, mean (±SEM) (n=6) | 97,3 (2,6) | - |
| Adaptation assessment score <70, No. (%) | 0 (0) | - |
| Adverse outcome (death, low IQ or cerebral palsy) | 6 (60) | - |
| Cerebral palsy, No. (%) (n=9) | 5 (55,5) | - |
| Low IQ (<85 Cognitive score), No. (%) (n=9) | 3 (33,3) | - |
| Epilepsy, No. (%) (n=9) | 3 (33,3) | - |

Abbreviations: PT, preterm infant; GM-IVH, germinal matrix-intraventricular hemorrhage; PHVD, post-hemorrhagic ventricular dilation; BPD, bronchopulmonary dysplasia; CRIB, Clinical Risk Index for Babies; TBV, Total brain volume; VV, Ventricular volume; IVD, right ventricular index; IVI, left ventricular index; AHWD, right anterior horn width; AHWI, left anterior horn width; TODD, right thalamus occipital distance; TODI, left thalamus occipital distance; IQ, intelligence quotient; cc, cubic centimeter; SEM, standard error of the mean; No., number; %, percentage; IQR, interquartile range.
\*Data only PT

### Specimen collection and processing

All CSF samples were acquired via taps from a ventricular reservoir in PT infants with PHVD or standard lumbar puncture in controls. CSF samples were collected by the neonatal intensive care unit clinical team at Puerta del Mar University Hospital using standard, sterile procedures. Reservoir taps were performed periodically with the aim of keeping the ventricular width below the 97th percentile. If taps were still necessary when the infant reached 1800g, a ventriculoperitoneal shunt was placed. CSF samples were sent to the clinical laboratory for clinically indicated microbiological culture (5-day surveillance) and laboratory evaluation. CSF aliquots were immediately frozen in sterile polypropylene tubes and stored at −80 °C until analysis. Samples were collected at different ages from birth (9-79 days) in PT with GM-IVH and PHVD as needed. CSF from control patients was collected from 3 to 28 days of age. Samples were centrifuged at 2000xg for 10 minutes at 20°C (or controlled room temperature) and the supernatant was divided into 150μL aliquots and stored at -80°C until use. CSF samples with blood were not used for biomarker studies. Only samples with a maximum of 2 freezing/thawing cycles were used in the study.

All CSF samples were obtained after parental consent in preterm patients with PHVD and in infants undergoing lumbar puncture for sepsis workup. The study was approved by Puerta del Mar University Hospital Ethics Committee in accordance with the Declaration of Helsinki.

### Neurodevelopmental assessment

At 2 years of age, a neurodevelopmental assessment was performed using the Bayley Scales of Infant and Toddler Development, third edition (BSID-III) (Bayley, 2006). Outcome was graded at 2 years of age as follows: (1) normal; (2) abnormal, i.e., children with a score of <70 in any of the 3 developmental domains on the BSID or cerebral palsy (Rosenbaum et al., 2007), and a level II-V of the Gross Motor Function Classification System (Palisano et al., 1997).

### Biochemical analysis

ELISA kits were used to analyze total tau (FRI99021,Fujirebio Europe N.V., Gent, Belgium), phosphor-Tau (pTau-S181) (FRI81361,Fujirebio Europe N.V., Gent, Belgium), neurofilament-light (NF-light) (10-7001, UmanDiagnostics AB, Umea, Sweden), phosphorylated neurofilament heavy (pNF-Heavy) (ELISA-pNF-H-V2, EnCor Biotechnology Inc., Gainesville, FL, USA), chitinase 3-like 1 (CHI3L1, also named YKL-40) (DC3L10, R&D Systems Inc, Minneapolis, MN, USA), myelin basic protein (MBP) (orb407124, Biorbyt Ltd., Cambridge, United Kingdom), SBDP120 (EH4242, Wuhan Fine Biotech Co., Ltd, Wuhan, China), Alpha II Spectrin Breakdown Product 145 (SBDP 145) (EH4243, Wuhan Fine Biotech Co., Ltd, Wuhan, China) and SBDP 150 (EH4244, Wuhan Fine Biotech Co., Ltd, Wuhan, China) following the manufacturer’s instructions. Absorbances were measured at 450nm on a spectrophotometer (MQX200R2, Bio-Tek Instruments, Burlington, VT, USA).

Ultra-sensitive Single-Molecule Array (SIMOA) technology on SR-X™ instrument (Quanterix, Inc., Billerica, MA, USA) was used to measure glial fibrillary acidic protein (GFAP) and ubiquitin carboxyl-terminal esterase L1 protein (UCHL1) from Simoa Human Neurology 4-Plex B assay (103345, Quanterix, Inc., Billerica, MA, USA) were quantified in CSF (1:40 dilution) following manufacturer’s instructions.

Cytokine/chemokine levels in CSF (1:2 dilution) were measured by the Bio-Plex Pro™ Human Cytokine 27-plex assay kit (M500KCAF0Y, Bio-Rad Laboratories, Inc., Hercules, CA, USA) that included the following markers: FGF basic, Eotaxin, G-CSF, GM-CSF, IFN-γ, IL-1β, IL-1ra, IL-2, IL-4, IL-5, IL-6, IL-7, IL-8, IL-9, IL-10, IL-12 (p70), IL-13, IL-15, IL-17A, IP-10, MCP-1 (MCAF), MIP-1α, MIP-1β, PDGF-BB, RANTES, TNF-α and VEGF. Assay was carried out on a Bio-Plex® 200 System (171000201, Bio-Rad Laboratories, Inc., Hercules, CA, USA) following strictly the manufacturer’s instructions. All samples in the study were run in duplicate on single-blinded study.

### Statistical analysis

All data are shown as mean ± standard error of mean (SEM). To quantify the relationship between different cellular and pathological markers and age, least squares linear regression models were used. Spearman’s correlation coefficient, ρ, together with the p-value were calculated and p < 0.05 was considered statistically significant. All analyses were performed using R.

### Hierarchical clustering

All data for each marker were z-scored across samples. Hierarchical clustering was performed using a Euclidian distance measure and Ward.D2 agglomeration via R.

### Partial least squares regression

Partial least squares regression (PLSR) analysis was used to analyze inflammatory cytokines and pathological markers. PLSR analyses find axes, called latent variables (LVs), which maximally identify the co-variance between the protein predictor variables and the outcome variable. LVs provides profiles of markers that are separated on samples based on continuous dependent variable (i.e., Age). PLRS analysis was performed in R using plsRglm package. Data were z-scored prior putting into the algorithm. To find maximal sample separation based on continuous dependent variable along the LVs axes, orthogonal rotations were applied to the sample scores and analyte weightings were computed using *opls* package in R.

### Linear Mixed Model for Repeated Measurements

We employed linear mixed model to compare cytokine expression and neurodegeneration marker levels between patients with motor assessment scores >70 (More) versus <70 (Less) using *limma* package available on Bioconductor in R. To account for repeated measurements within each patient, we first estimated the within-patient (i.e., multiple samples can share same patient ID) correlation using the *duplicateCorrelation()* function. Then, these correlation values were specified in the *correlation* argument of the *lmFit()* function. Patient ID was included as the *block* variable to indicate repeated samples originating from the same patient. By incorporating both the block variable and within-patient correlation, the model accounted for dependence of repeated measures. The design matrix included Motor Assessment group (reference = Less) and Age as fixed effects. Lastly, p values were adjusted for false discovery rate (FDR) across multiple measurements (i.e., Cytokines, Neurodegeneration markers) using the Benjamini-Hochberg (α=0.05).

## RESULTS

### Neuropathological decline with age

The aim of this work is to illuminate temporal relationships between function and pathological changes with age in preterm patients. To do so, we sampled functional scores, ventricular volume, and CSF at defined time points from birth until discharge or death (**Fig. 1A, Materials and Methods, Table 1, S1**), then quantified a panel of neuropathological markers, including NF-light, phosphorylated tau (pTau S181), and glial fibrillary acidic protein (GFAP) (**Fig. 1B**). Enlarged ventricles are commonly observed in PT infants and are considered key indicators of prematurity and associated periventricular tissue injury (Shi et al., 2015). Here, we did not identify significant changes in ventricular volume with age (**Fig. 1C**). Although, previous research has reported reduced brain volumes and enlarged ventricular volumes in PT a at term equivalent age (Brouwer et al., 2012) and remained when they age to adolescence (Nosarti et al., 2002), our analysis is limited to the first two months of life (up to 60 days post-delivery). Therefore, we cannot rule out the possibility of long-term alterations in our patients. We also assessed GFAP levels, a putative marker of reactive astrogliosis and brain damage, which increases in CSF within the first 48 hours, following acute brain injury (Blennow et al., 1996). Our temporal analysis of GFAP showed a trending decrease with age (**Fig. 1D**).

**Figure 1.**
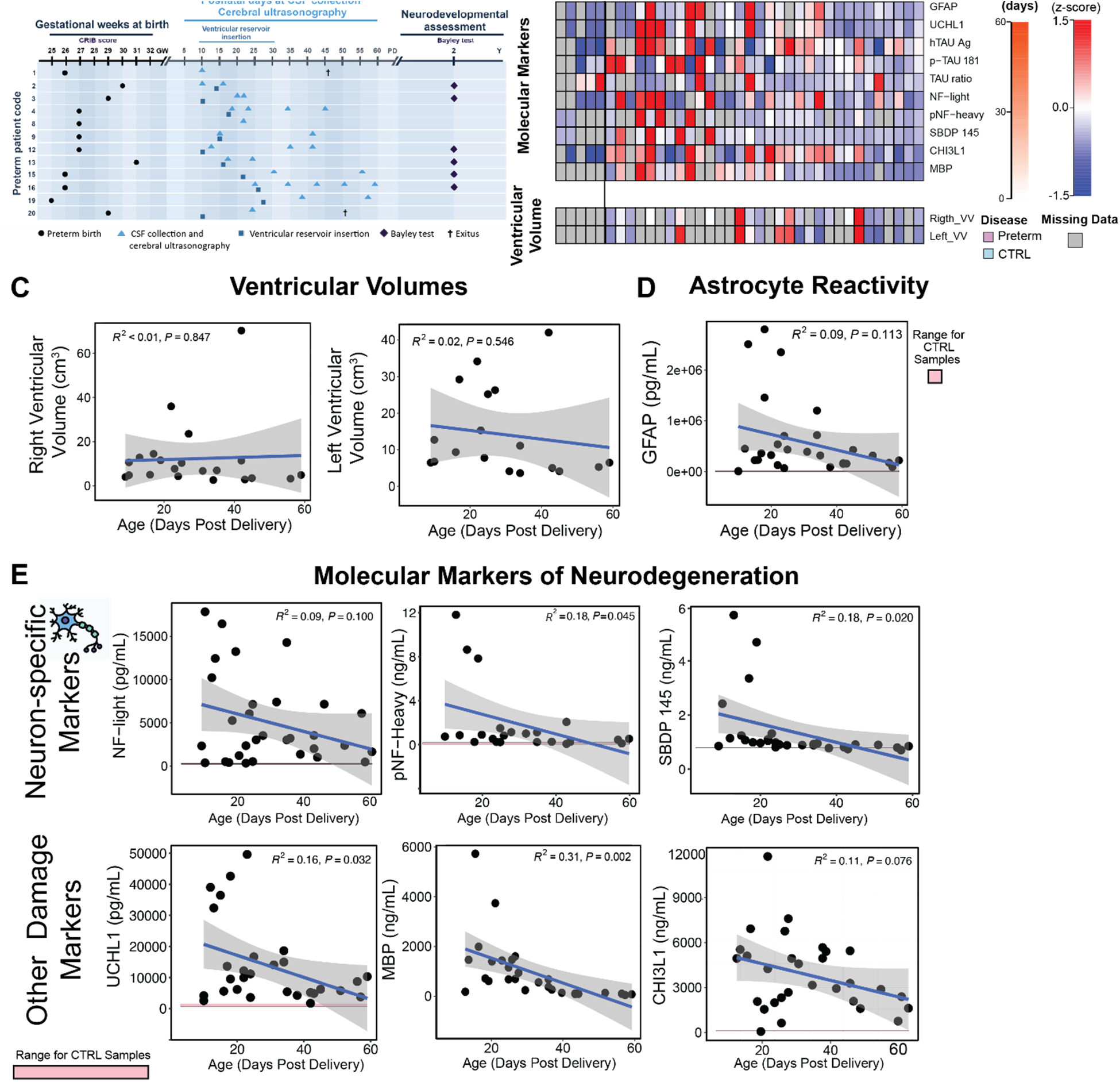
Neurodegenerative markers decline with age in PT patients. (**A**) Schematic of data collection from PT patients. Markers: death, triangle: sequential, square: shunt placed. (**B**) Neurodegenerative and neuropathological measures sorted by age (data are z-scored, missing data are colored in grey). (**C**) ventricular volume vs. age. (**D**) Astrocyte reactivity marker GFAP quantified in CSF vs. age (grey area indicates 95%CI from linear regression, p-value from Spearman correlation). (**E**) Molecular neurodegenerative markers quantified in CSF vs age (pink boxes show range of control patients, grey area represents the 95% confidence interval for the linear regression, blue line denotes the fitted regression, correlation coefficient R and corresponding p value are shown.)

To assess neuronal damage, we also quantified neuron-specific injury markers in the CSF, including neurofilaments and tau protein **(Fig S1)**. Neurofilaments, which are proteins of the neuronal scaffold, are released into the CSF when acute or chronic damage occurs (Goeral et al., 2021). We assessed the subunits corresponding to NF-light and pNF-Heavy chain proteins, which showed a decrease toward the range of healthy controls in the levels of both markers with time (**Fig. 1E**). SBDP 145, as marker of cytoskeletal alterations and neuronal damage (Yan & Jeromin, 2012), also showed a decrease toward control values as time post-delivery progressed (**Fig. 1E**). We also evaluated other markers of general brain injury (**Fig. S1**). CHI3L1 plays a key role in inflammation and tissue remodeling in response to injury (Ijabi et al., 2024), and showed a trending toward to control values over time (**Fig. 1E**). Additionally, UCHL1 is involved in the maintenance of axonal integrity and synaptic function as well as in the inflammatory response (Matuszczak, Tylicka, Komarowska, Debek, & Hermanowicz, 2020), and MBP is a component of the myelin sheath surrounding axons, that serves as a marker of neuronal and white matter damage severity (García-Alix et al., 1994). These markers were decreasing toward the range of healthy controls with time (**Fig. 1E**). These data thus suggest that our PT cohort consists of early injury with degeneration markers that decline with time.

### Longitudinal analysis of CSF cytokine levels in PT patients identifies groups of transient and sustained cytokines

Having established that pathological and functional markers, including GFAP, improve with age, we next asked if the CSF inflammatory milieu would also subside with age. To test this, we quantified a panel of 27 cytokines in the CSF via a Luminex multiplexed immunoassay (**Figs. 2A, S2-3**). Hierarchical clustering revealed that there were two distinctive cytokine groups. The first consisted of cytokines that remained sustained with age, while the second consisted of cytokines that diminished with age (**Fig. 2B, C**). Individual analysis of sustained cytokines, including pro-inflammatory cytokines IFN-γ, IL-7 and MCP-1, remained elevated and did not return to baseline control levels within the observation window (**Fig. 2B**). Conversely, diminishing cytokines, including pro-inflammatory cytokines like IL-8, IL-9, IP-10, MIP-1β and IL-6, were individually significantly correlated with age and declined to baseline levels with time (**Fig. 2C**). These distinctive responses indicate that certain aspects of the immune response decrease with time, while others do not.

**Figure 2.**
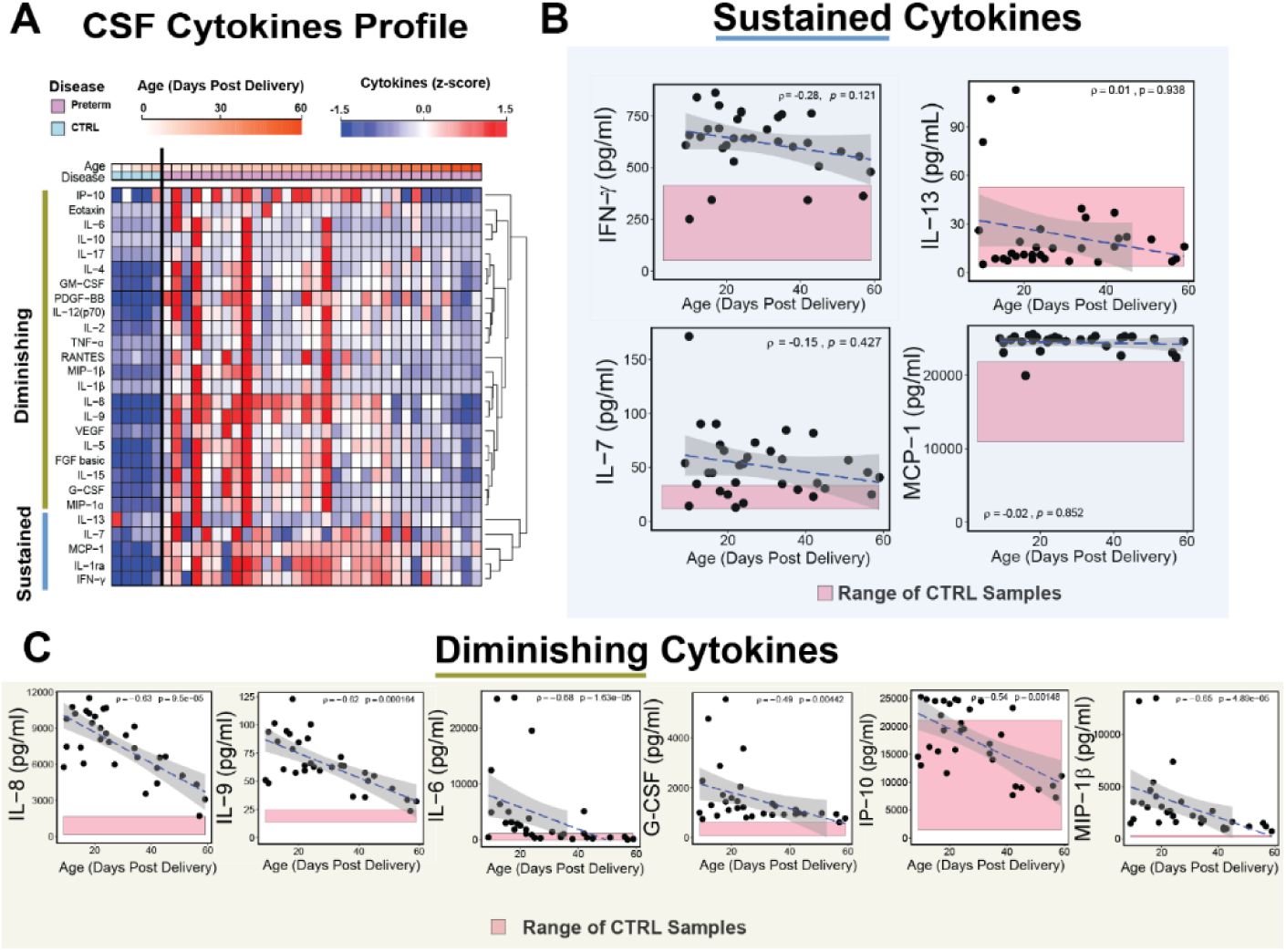
Longitudinal analysis of CSF cytokine levels identifies profiles of diminishing and sustained levels. (**A**) Heatmap representation of key neuroinflammatory cytokines is shown (All data z-scored). CSF cytokine profiles elicited two distinct expression patterns over time. (**B**) Sustained cytokines vs. age (pink boxes show range of control patients, grey indicates 95%CI from linear regression, p-value from Spearman correlation). (**C**) Diminishing cytokines vs. age (pink boxes show range of control patients, grey indicates 95%CI from linear regression, p-value from Spearman correlation).

### Sustained cytokines are associated with GFAP

Having found that both GFAP and certain cytokines were sustained across time (e.g., MCP-1, IL-8), we next asked if certain cytokines would be related to GFAP levels in the CSF. To account for the multidimensional nature of the data, we used a partial least squares regression (PLSR) on log2 transformed GFAP values (achieving normally distributed data, **Fig. S4**) to identify a profile of cytokines correlated with GFAP. The analysis separated high GFAP values to the right and low GFAP values to the left along latent variable 1 (LV1) (**Fig. 3A**). LV1 consisted of a profile of cytokines that correlated with higher GFAP (positive) or lower GFAP (negative). Top cytokines correlated with GFAP levels include several that were sustained, including MCP-1, IFN-g and IL-1ra, and several that were diminishing with age, such as IL-8 and MIP-1α (**Fig. 3A, B**). These top correlating cytokines, mainly include pro-inflammatory chemokines, such as IL-8 and MCP-1, together with IFN-g, which are all highly secreted by reactive astrocytes (Folkerth et al., 2004; Robinson, Narasipura, Wallace, Ritz, & Al-Harthi, 2020; Singh, Anshita, & Ravichandiran, 2021; Van Der Voorn et al., 1999). Interestingly, IL1-ra was also a top correlate, which plays a fundamental role in modulating the inflammatory response (Arend, Malyak, Guthridge, & Gabay, 1998). Conversely, down-regulated cytokines correlated with lower GFAP levels, include Eotaxin, an eosinophil chemoattractant cytokine (Raba & Tabarkiewicz, 2018) and IL-6 and IL-13, with both pro- and anti-inflammatory effects (McAdams & Juul, 2012) (**Fig. 3A**).

**Figure 3.**
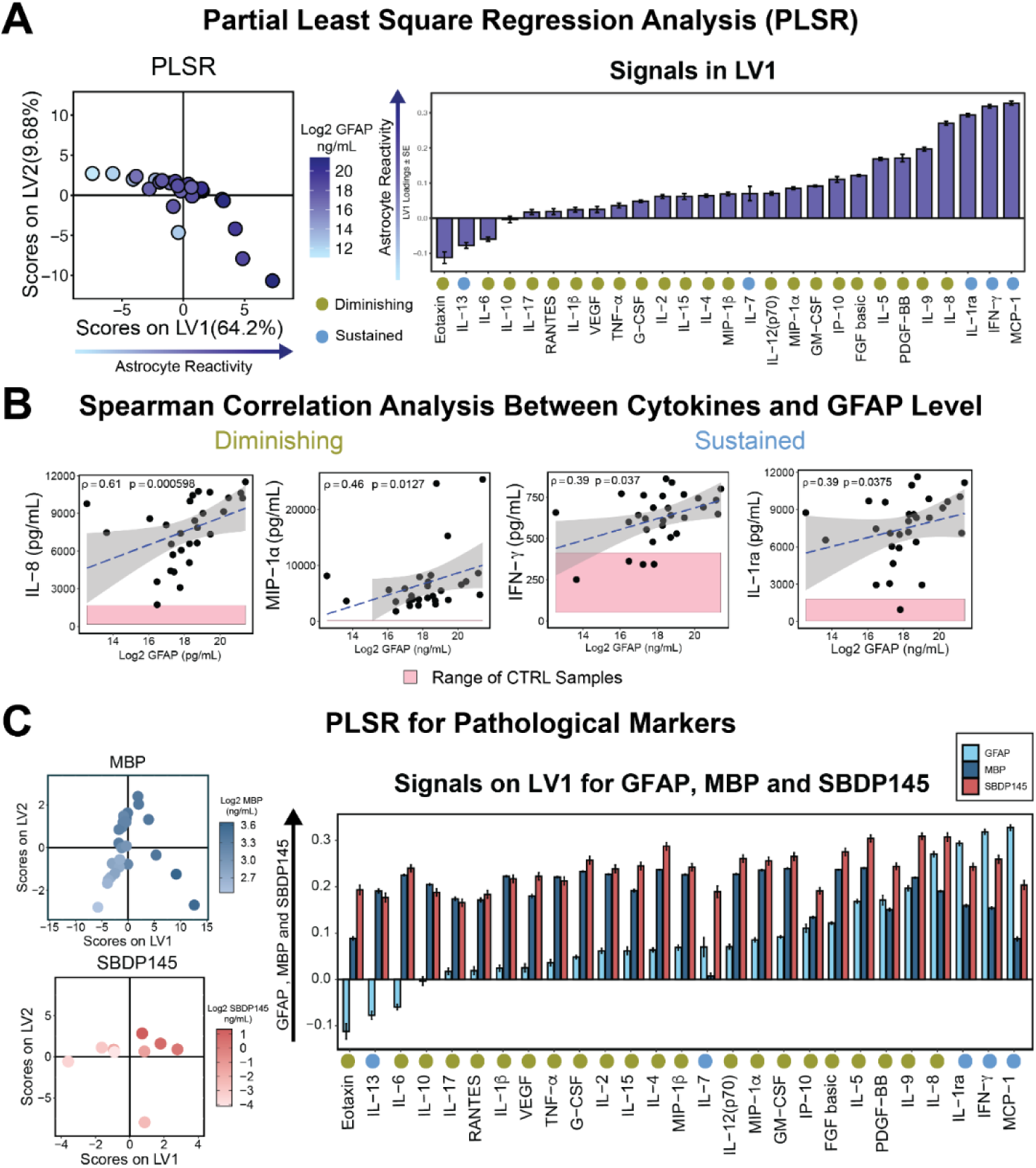
Pro-inflammatory cytokines correlate with GFAP in the CSF. (**A**) PLSR of cytokines vs GFAP identified an axis, latent variable 1 (LV1) that separated higher GFAP levels to the right and lower GFAP levels to the left. LV1 consisted of a profile of cytokines correlated with GFAP (positive) or inversely correlated (negative) (error bars mean ± SEM). (**B**) The positively correlated cytokines included both diminishing, such as IL-8, and sustained cytokines, such as IL-1ra, MCP-1 and IFN-γ (pink boxes show range of control patients, grey indicates 95%CI from linear regression, p-value from Spearman correlation) (**C**) Conducting the same PLSR of cytokines against MBP and SBDP145 revealed distinctive profiles of cytokines correlated with each. (Blue circles correspond to sustained cytokines, green circles correspond to diminishing cytokines).

Taking into account that high levels of GFAP seem to correlate with astrogliosis and brain damage, mainly in periventricular white matter injury in PT (U. Sjöbom et al., 2021; Stewart et al., 2013; Whitelaw, Rosengren, & Blennow, 2001), we used a PLSR on log2 transformed to analyze the cytokine profile of other pathological markers such as MBP, characteristic of white matter damage, and SBDP 145, marker of cytoskeletal alterations and neuronal damage (Yan & Jeromin, 2012). The cytokine profile observed in LV1 for GFAP, MBP and SBDP145, showed a differential pattern observed (**Fig. 3C**). These findings suggest that reactive astrocytes may promote a pro-inflammatory environment or might be overexpressed as a consequence.

### White Matter Lesions Correlate with Numerous Cytokines

Because the multivariate PLSR analysis showed that cytokines strongly correlated with both GFAP and markers of glial damage, we next asked if individual cytokines would strongly correlate with individual markers of neuronal or white matter damage. To do so, we conducted individual correlation analyses between each cytokine and all damage markers (**Figs. 4A, Table S2**). We found that robustly significant correlations between many of the cytokines and CSF levels of SBDP145 and MBP. Interestingly, certain cytokines, including IL-6, IL-10, IP-10, and IL-17 were significantly correlated with ventricular volume (Gongvatana et al., 2014; Schmidt et al., 2016), suggesting that these cytokines are directly related to gross physiological damage (**Fig. 4B**). Specific cytokines are also significantly correlated with CHI3L1, MPB, and SBDP145 (**Fig. 4C**), suggesting that some of these could be preferentially driven by neuronal or white matter damage. CHI3L1 exerts pro-inflammatory activity regulated by IL-6 (Liu, Hu, Ding, Li, & Ding, 2024) and correlates with IL-2, which has been implicated in perinatal white matter injury (Kadhim, Tabarki, De Prez, Rona, & Sebire, 2002). Notably, CHI3L1 also correlates with IL-4, a predominantly anti-inflammatory cytokine, which at high concentrations may have adverse effects on PT development (Alan Leviton et al., 2018; Opal & DePalo, 2000). Also, the analysis of cytokines correlated with MBP and SBDP145 revealed positive correlations with MIP-1β, IL-8 and IL-6, all linked with white matter damage and neuritic damage in PT (Ellison et al., 2005; A. Leviton et al., 2013; Sullivan et al., 2020).

**Figure 4:**
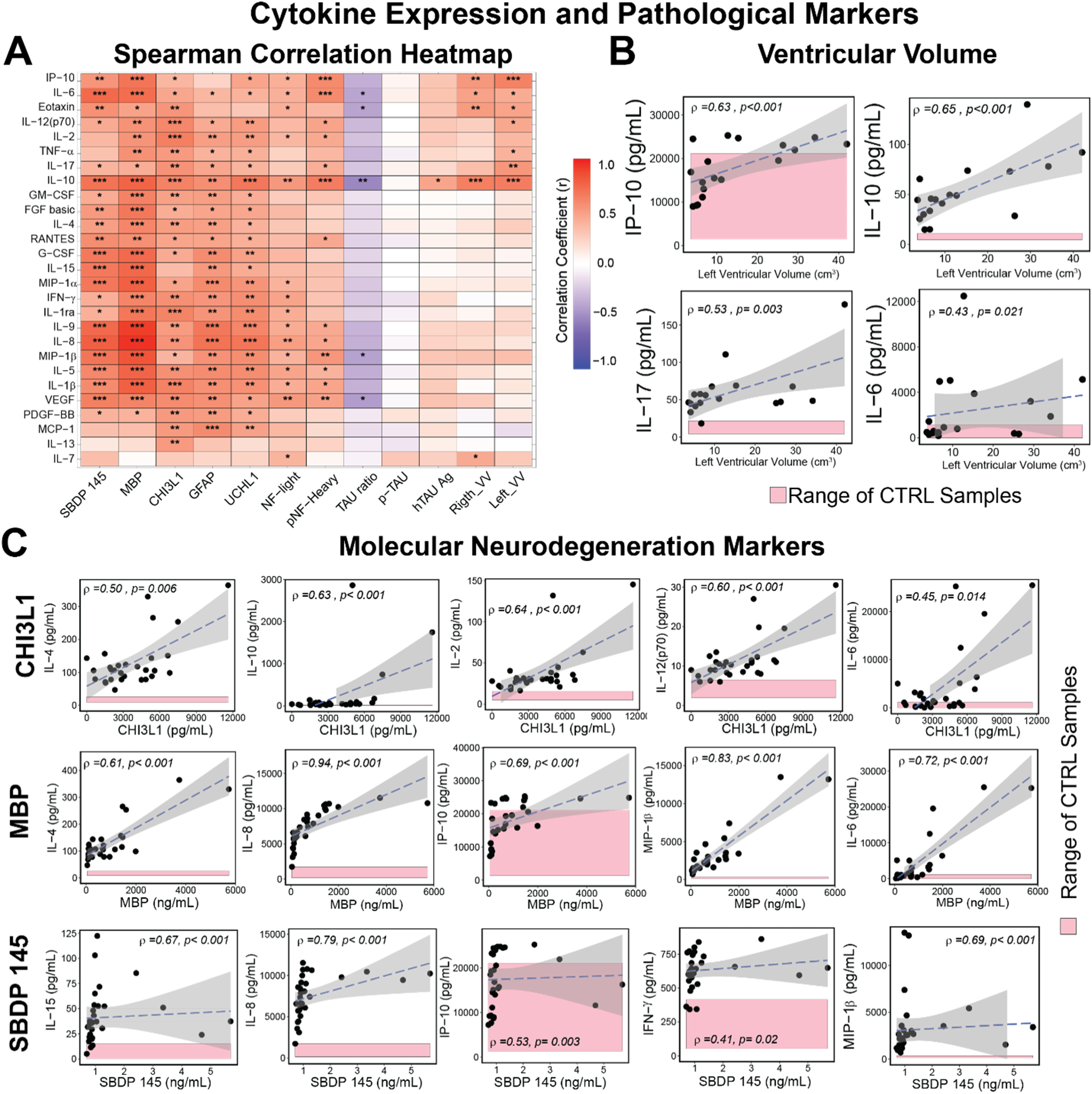
Cytokines significantly correlate with molecular neurodegeneration markers and ventricular volume. Heatmap of spearman correlation coefficient of cytokines and pathological and neurodegeneration markers. The color bar represents correlation coefficient where negative - 1 represents strong negative correlation (dark blue) and +1 represents strong positive correlation (dark red). Significance levels (FDR adjusted p-value) are indicated as asterisk (*p < 0.05, **p<0.01, ***p<0.001, and ****p<0.0001. (**B-C**) Spearman correlation of cytokines with ventricular volume and molecular neurodegeneration markers including CHI3L1, MBP and SBDP145. (Pink boxes show range of control patients, grey indicates 95%CI from linear regression, p-value from Spearman correlation).

### Pathological Cytokines are associated with cognitive and motor outcomes

We concluded this work by asking if any of the CSF cytokines or neurodegeneration markers quantified in the CSF within the first 60 days of life would relate to long term functional outcome measures assessed at >2 years. In this study, we possessed paired molecular and functional data from 6 patients spanning multiple time points (**Table 1**). Correlations between cytokines and clinical measures revealed that specific acute cytokines, including VEGF, Eotaxin, IL-6 were significantly correlated with cognition and adaptation, while IL-7 was significantly negatively correlated with motor assessment and social emotional assessment (**Fig. 5A, Table S2**). Because repeated molecular measures were used for each sample, we next evaluated if we could identify significant differences in molecular markers, when patients were grouped based on better or poorer longer-term functional outcomes (**Table S3**). We grouped patients based on motor assessment score into Low (<70) or High (>70) attending to previous considerations (Rosenbaum et al., 2007). Because we had multiple molecular CSF measurements for each patient, we accounted for the repeated measures using a linear mixed effects model that included terms for motor function group (Low vs High) and patient age when the sample was collected. This analysis identified significance associated with motor function group in CHI3L1 (**Fig. 5B**) and several sustained cytokines (**Fig. 5C**). Although based on a relatively small sample size, these data suggest a potential relationship between higher levels of sustained cytokines and poorer longer-term outcomes.

**Figure 5:**
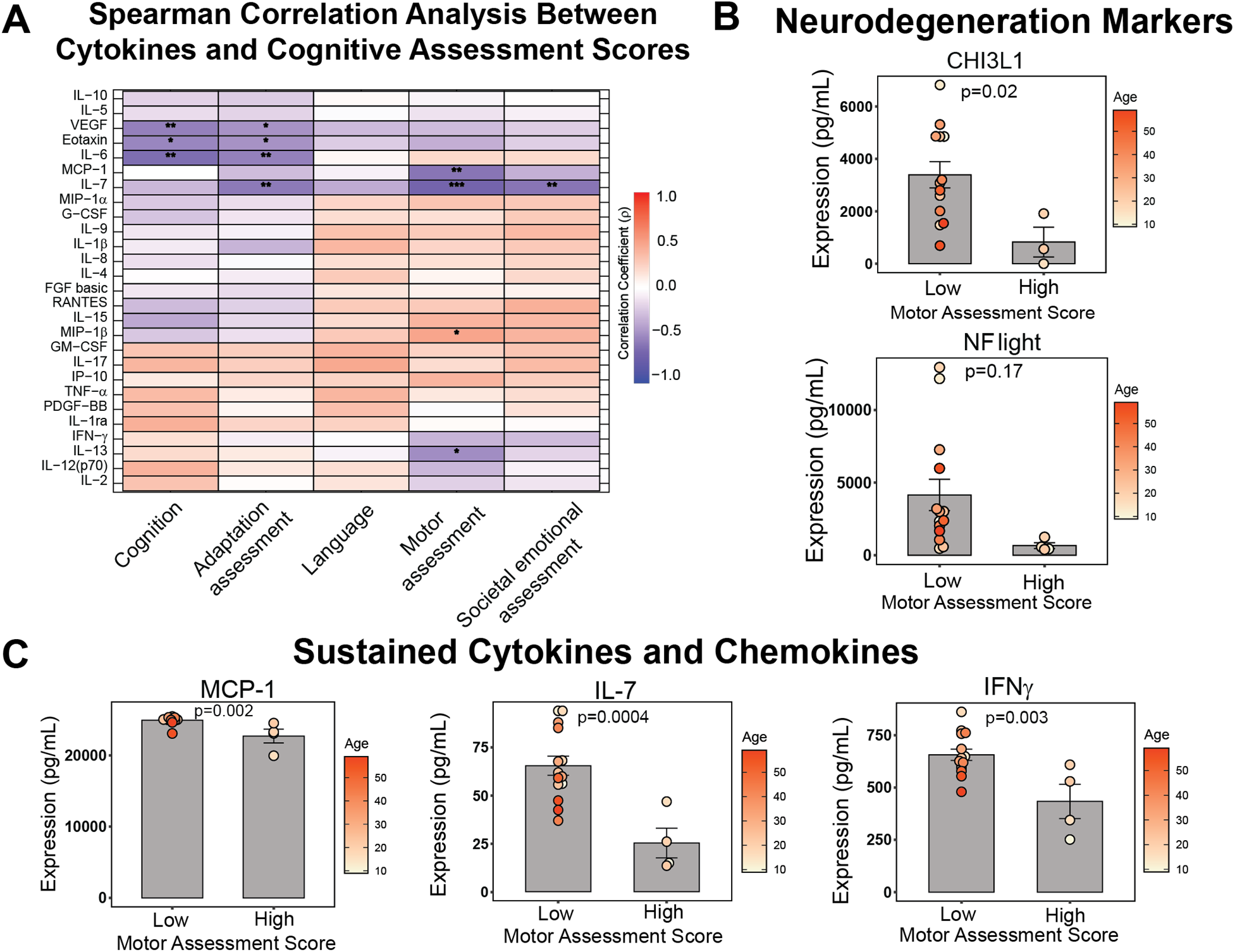
Early sustained cytokines and neurodegeneration markers are associated with long term motor function deficit. (**A**) Inflammatory cytokines are highly correlated with behavioral and cognitive assessments. Significance levels (FDR adjusted p-value) are indicated as asterisk (*p < 0.05, **p<0.01, and ***p<0.001) (**B**) Patients with lower CHI3L1 levels have higher motor score in the long term (>2 yrs) (N=4-6 for each group, statistical analysis computed via linear mixed model, mean ± SEM). (**C**) Long term motor assessment score is related to early sustained cytokine level (N=4-6 for each group, statistical analysis computed via linear mixed model, mean ± SEM). Bars represent children with BSID-III motor scores below 70 (Low) and above 70 (High).

## DISCUSSION

In this study, we examined longitudinal changes in the cytokine profile and in glial and neuronal markers in the CSF of 12 PT infants with very low birth weight who presented with grades III-IV GM-IVH and PHVD. PHVD develops in up to 50% of preterm infants with severe GM-IVH (Murphy et al., 2002), typically requiring shunt placement (de Vries et al., 2002; Leijser et al., 2018), and is associated with a worse prognosis and poorer neurodevelopmental outcomes (Benavente-Fernández, Steggerda, Liem, Lubián-López, & de Vries, 2023; Leijser et al., 2018; Maunu et al., 2011; Srinivasakumar et al., 2013). We therefore hypothesized that levels of damage markers and cytokines would relate to both time after IVH and longer-term functional outcomes. To our knowledge, this is the first study to evaluate longitudinal changes in this comprehensive panel of injury markers and cytokines in the CSF and related them to longer term functional outcomes.

Neurofilament proteins, such as NF-light, are increasingly recognized as highly sensitive and robust biomarkers of neuroaxonal damage and neurodegeneration in neurological diseases like multiple sclerosis, Alzheimer’s disease and traumatic brain injury, among others, being released following axonal disruption (for review (Khalil et al., 2024; Yuan & Nixon, 2021)). Elevated blood NF-light concentrations at specific times after birth have been reported in PT infants and correlate with indices of brain maturation, severity of injury, gestational age, and birth weight (Depoorter et al., 2018; Ulrika Sjöbom et al., 2025). Our longitudinal analysis shows that NF-light levels declined over time, although values remained elevated. Consistently, increased CSF NF-light levels have been observed up to 12 months of age in preterm infants with GM-IVH and PHVD with parenchymal lesions, particularly among those with neurodevelopmental disability (Ruth Griffiths developmental quotient <70) (Whitelaw et al., 2001). Furthermore, serum NF-light concentrations remain elevated in PT for up to one month after birth (U. Sjöbom et al., 2021).

We profiled a broader battery of cellular injury markers, including SBDP-145, a proteolytic fragment of the neuronal cytoskeletal protein αII-spectrin, generated in response to axonal injury (Pineda et al., 2007; Pritt et al., 2014). SBDP-145 concentrations were highest in the early days following IVH–MG–PHVD and decreased progressively thereafter. These findings are in agreement with prior work reporting elevated SBDP-145 levels in PT with GM-IVH and white matter injury compared with term controls, with concentrations correlating with the severity and extent of brain injury (S. Cheng, Sun, Li, & Dong, 2024). Moreover, SBDP-145 has been proposed as a potential biomarker of lesion severity and prognosis in both traumatic brain injury and subarachnoid hemorrhage (Papa et al., 2018; Sari, Oswari, Ong, Adam, & Atik, 2025).

White matter damage is also commonly observed in PT patients and affects up to 50% of preterm infants with MG-IVH (Ballabh & de Vries, 2021; Volpe, 2003). Interestingly, when PHVD is observed in PT with GM-IVH, as observed in our study cohort, it is associated with structural alterations in commissural nerve fibers located adjacent to the dilated ventricles (Nieuwets et al., 2022) and up to 78% of PT with severe GM–IVH and PHVD exhibit varying degrees of white matter injury (Ballabh & de Vries, 2021). Here, we quantified CSF MBP levels as a marker of white matter damage and neuronal injury secondary to demyelination (Polis, Polis, Zeman, Paśnik, & Nowosławska, 2020). Elevated CSF MBP concentrations have been described in several neurodegenerative and cerebrovascular disorders, including multiple sclerosis and stroke (Agoston, Shutes-David, & Peskind, 2017; Lamers et al., 1995; Sellebjerg et al., 2017), as well as in neonates with hypoxic–ischemic encephalopathy (García-Alix et al., 1994). In our cohort, MBP levels decreased progressively over time. To our knowledge, no previous study has assessed the levels of MBP in CSF from PT, and MBP plasma levels are higher in PT with white matter damage (Zhao et al., 2022), and MBP levels seem to increase up to 3 days after birth in patients with periventricular-IVH (Zhou et al., 2015) The temporal pattern we observed, characterized by a gradual decline in MBP concentrations, likely reflects the resolution of acute white matter injury, although the structural lesions established during the initial phase may persist beyond the period of biochemical normalization.

The damage observed in PT is also associated with astrocyte dysfunction, characterized by their reactive transformation and the development of astrogliosis in response to injury (L. Li et al., 2025; Mallard et al., 2014; Van Steenwinckel et al., 2024; Wisnowski et al., 2014). In addition, PT with white matter injury (Agut et al., 2020) or MG-IVH with PHVD (El-Dib et al., 2020) frequently present gliosis (Ballabh & de Vries, 2021). We therefore quantified GFAP as a specific marker of reactive astrocytes and is upregulated during astrogliosis (Agnello et al., 2025; Blennow et al., 1996; Eng & Ghirnikar, 1994; L. Li et al., 2025). We found that GFAP tends to decrease with time. GFAP is known to be elevated in both serum and CSF in association with white matter injury and brain damage (U. Sjöbom et al., 2021; Stewart et al., 2013). However, serum and CSF GFAP concentrations do not appear to depend on gestational age (Blennow et al., 1996; U. Sjöbom et al., 2021). Also, increased CSF GFAP levels are observed in PT with PHVD who also present white matter injury (Whitelaw et al., 2001). Furthermore, reactive astrocytes secrete both pro- and anti-inflammatory mediators, including IL-8, IL-5, and CHI3L1 (Kwon & Koh, 2020). Among these, CHI3L1 is released by astrocytes in response to proinflammatory cytokines derived from macrophages and is associated with astrogliosis (Bonneh-Barkay et al., 2012; Connolly et al., 2023). Similar to GFAP, we found a trend to decrease in CHI3L1 levels in our cohort. Nevertheless, CHI3L1 levels are increased in CSF and plasma in several neurodegenerative diseases with neuroinflammatory profile (Bonneh-Barkay, Wang, Starkey, Hamilton, & Wiley, 2010; Jiang et al., 2023; Llorens et al., 2017; Pase et al., 2024; Yu et al., 2024). Despite its extensive use as a biomarker of neuronal injury and inflammation in neurodegenerative conditions, CHI3L1 has not yet been studied in the context of brain injury in PT. Notably, because CHI3L1 can also be released by other cell types, including neutrophils, macrophages, and epithelial cells (Yu et al., 2024), it has been used as a biomarker for bronchopulmonary dysplasia (Henckel et al., 2021; James et al., 2021; König et al., 2016), another pathology associated with prematurity. Although we observed trend to decrease in both GFAP and CHI3L1 levels, these proteins are well-established markers of gliosis and inflammation and are typically upregulated by proinflammatory stimuli (D. Zhang, Hu, Qian, O’Callaghan, & Hong, 2010; W. Zhang et al., 2023). These findings may suggest the presence of a highly pronounced, and a persistent inflammatory environment in the brains of these PT.

Taking these findings into account, we analyzed the temporal profile of cytokines present in the CSF of our PT. It is important to note that the neonatal immune system is immature, and this immaturity is even more pronounced in PT (Melville & Moss, 2013), and their cytokine profiles may differ substantially from those of full-term neonates (Boardman et al., 2018).

Nevertheless, their capacity to mature and mount responses to injury or infection does not appear to be impaired (Kamdar et al., 2020). Longitudinal studies have assessed CSF levels of IL-6, IL-8, IL-10, TNF-α, and IFN-γ in PT over a period of up to 121 days, without detecting significant temporal changes (Ellison et al., 2005). However, this study did not account for the potential influence of prematurity-related complications. In contrast, based on the temporal patterns observed in our cohort, cytokines could be broadly categorized into two groups: (i) those that exhibited a significant temporal decrease within the observation period (including IL-8, IL-9, IL-6, IP-10, G-CSF, and MIP-1β, among others), and (ii) those with sustained levels over time (IFN-γ, IL-13, IL-7, IL-1rα, and MCP-1). Among decreased cytokines, IL-6 and IL-8 are well known mediators of inflammatory response associated with preterm delivery and PT brain injury (Holst, Mattsby-Baltzer, Wennerholm, Hagberg, & Jacobsson, 2005; Jung et al., 2016; Schultz et al., 2002; Yoon et al., 1996). IL-6 is a pleiotropic cytokine that presents a dual pro- and anti-inflammatory effect in the CNS (Biber et al., 2008; Erta, Quintana, & Hidalgo, 2012; Grebenciucova & VanHaerents, 2023; Kummer, Zeidler, Kalpachidou, & Kress, 2021). IL-6 is also an essential player for the establishment of chronic inflammation (Gabay, 2006; Prairie et al., 2021) and disruption of BBB (Yang et al., 2022). Similarly, IL-8 is a pro-inflammatory cytokine, linked to brain dysmaturation in PT (Kossmann et al., 1997; Sullivan et al., 2020). Thus, CSF IL-6 and IL-8 are found to be increased at 72h in PT with hypoxic-ischemic encephalopathy (Karin Sävman, Blennow, Gustafson, Tarkowski, & Hagberg, 1998) and remained elevated two weeks after birth in PT with PHVD (K. Sävman et al., 2002). However, IL-6 is only increased in PT with posthemorrhagic hydrocephalus (Habiyaremye et al., 2017) or in patients with white matter injury (Ellison et al., 2005). Also, IP-10, a chemokine secreted in response to IFN-γ and associated with BBB disruption (Madhurantakam, Lee, Naqvi, & Prasad, 2023; K. Wang et al., 2018), was elevated in PT with posthemorrhagic hydrocephalus at two weeks of age (Habiyaremye et al., 2017) and IL-9 and MIP-1β, involved in excitotoxicity and BBB disruption (Boardman et al., 2018; Donninelli et al., 2020) were higher in PT compared with term infants (Boardman et al., 2018).

Among the cytokines with sustained levels, IFN-γ and MCP-1 concentrations remained stable throughout the observation period. Previous studies have reported no significant differences in CSF IFN-γ and MCP-1 levels at approximately two weeks of age when comparing PT and full-term infants (Boardman et al., 2018; K. Sävman et al., 2002). Similarly, IFN-γ levels appear unaltered in PT with posthemorrhagic hydrocephalus up to two months of age (Habiyaremye et al., 2017; Schmitz et al., 2007), and MCP-1 levels remain unaffected at two weeks (Habiyaremye et al., 2017). In contrast, transient elevations of IFN-γ have been observed in PT with white matter injury (Schmitz et al., 2007), suggesting that disease-specific factors and the timing of sampling critically influence cytokine dynamics. However, bearing in mind that IFN-γ is a pro-inflammatory cytokine associated with white matter damage (Hansen-Pupp et al., 2005; Popko, Corbin, Baerwald, Dupree, & Garcia, 1997), and MCP-1 is a chemokine involved in the recruitment of monocytes and lymphocytes during inflammation process (Yao & Tsirka, 2014), may suggest greater severity of the damage in our PT cohort that could be responsible for both markers being elevated within our observation window.

When evaluating neurodevelopmental outcomes at two years of age using the Bayley Scales of Infant Development, and defining developmental delay as a score below 70 (Ballot et al., 2017; Rosenbaum et al., 2007), we found that lower motor scores (<70) were associated with higher concentrations of the sustained cytokines IFN-γ, MCP-1, and IL-7. The relationship between IFN-γ and neurodevelopmental outcomes remains controversial: while some studies have reported no association in PT at two years of age (Hansen-Pupp et al., 2008), others have shown that elevated serum IFN-γ levels are linked to neurodevelopmental impairment in infants born at 34 gestational weeks at one year of age (Wu et al., 2025). Similarly, increased blood MCP-1 levels have been associated with poorer neurodevelopmental outcomes and cerebral palsy (Nist & Pickler, 2019). Elevated plasma MCP-1 concentrations in transfused preterm infants born before 22 weeks of gestation have also been correlated with lower cognitive and motor Bayley scores at one year of age (Benavides et al., 2022). Conversely, higher serum IL-7 levels have been associated with better neurodevelopmental outcomes at one year (Wu et al., 2025), in contrast to our findings.

We also observed a trend toward higher CSF concentrations of NF-light, a well-established marker of neuronal injury, being associated with poorer motor scores at two years of age. This observation is consistent with previous studies reporting that elevated serum NF-light levels correlate with adverse neurodevelopmental outcomes at two years, particularly affecting motor performance at both one and 2 years of age (Goeral et al., 2021; U. Sjöbom et al., 2021). In parallel, increased levels of the neuroinflammation marker CHI3L1 were also associated with poorer motor outcomes. Although data on CHI3L1 in PT are lacking, this biomarker has been closely linked to white matter injury (Peterson et al., 2025; W. Zhang et al., 2023), which in turn is strongly associated with adverse motor development and cerebral palsy (Imamura et al., 2013; Woodward, Clark, Bora, & Inder, 2012). Together, these findings underscore the potential of neurodegenerative and inflammatory biomarkers to identify preterm infants at risk for adverse neurodevelopmental outcomes.

While our study longitudinally analyzed CSF in PT infants from birth to two months of age, providing a relevant window into the evolution of GM–IVH and PHVD pathology, as well as neurobehavioral assessment at two years of age, several limitations must be acknowledged. First, the sample size was relatively small, including only 12 PT for CSF studies and 6 infants available for Bayley developmental assessment, which warrants cautious interpretation of the associations identified. Second, due to the retrospective design, some morphometric data related to brain volume measurements were lost during data retrieval. In addition, CSF samples were obtained at different time points depending on clinical indications, which may introduce variability in the temporal analysis, although they still provide a meaningful representation of the pathological evolution. The control group also consisted of infants undergoing lumbar puncture for sepsis evaluation rather than healthy PT controls, which may limit direct comparisons between groups. Nevertheless, given the study population and sample type, control patients inevitably had to be selected from individuals undergoing evaluation for other pathological conditions.Also, the limited sample size warrant cautious interpretation of the associations identified. However, the comprehensive assessment of neuronal and neuroinflammatory markers in PT with IVH–MG and PHVD provides a more complete picture than previous studies that focused on single or a handful of biomarkers. Furthermore, we perform our analysis on CSF samples, which allows us to directly assess the immune environment surrounding the central nervous system of these children, unlike studies using peripheral samples such as blood or plasma. Given the limited number of studies investigating the temporal dynamics of neuroinflammatory responses in this population, our findings may serve as a foundation for identifying potential biomarkers of brain injury and guiding the development of new therapeutic strategies.

In summary, our longitudinal analysis of CSF in PT with GM-IVH for the first time identifies distinct profiles of inflammatory cytokines and neurodegenerative markers that either transiently decay or are sustained through the early postnatal period. Interestingly, our finding that sustained cytokines were associated with markers of astrogliosis and with poorer long-term motor outcomes and these pathways may warrant further investigation as potential therapeutic targets. Whereas further studies in larger cohorts are needed to confirm these associations, the longitudinal design of the study, the direct analysis of CSF biomarkers, and the integration of molecular findings with neurodevelopmental assessment provide new insights into the evolution of neuroinflammation and following GM-IVH in PT.

## Data Availability

All data produced in the present work are contained in the manuscript

## SUPPLEMENTARY TABLE CAPTIONS

**Supplementary Table S1**: Cytokine, pathology, functional, and metadata for all patient simples included in Study.

**Supplementary Table S2**: Correlation analysis between cytokines and pathology or functional outcomes.

**Supplementary Table S3**: Linear mixed model analysis of cytokines and neurodegeneration markers with long term functional déficits.

