## Supplementary Information for "Temporal Co-Evolution of Clinical Scores and Neuro-Immune Biomarkers in Preterm Infants with Severe Germinal Matrix-Intraventricular Hemorrhage and Post-Hemorrhagic Ventricular Dilation"

[2]Division of Physiology. School of Medicine, Universidad de Cadiz. Cadiz, Spain.

[3]Biomedical Research and Innovation Institute of Cadiz (INiBICA) Research Unit, Puerta del Mar University, Cadiz, Spain.

[4] Division of Neonatology, Department of Pediatrics, Puerta del Mar University Hospital, Cádiz, Spain

[5]Area of Pediatrics, Department of Child and Mother Health and Radiology, Medical School, University of Cadiz, Cadiz, Spain.

[6] Neonatal Unit, Hospital Universitario de Burgos, Burgos, Spain.

[7] Wallace H. Coulter Department of Biomedical Engineering, Georgia Institute of Technology, Atlanta, GA 30332 USA

**Running title:** Cytokines and neuropathological markers alleviate with time in Preterm Infants

[a]Equally contributing first authors

*Equally contributing senior authors.

*Address correspondence to:

| Levi B. Wood<br>Parker H. Petit Institute for Bioengineering & Bioscience<br>George W. Woodruff School of Mechanical Engineering<br>Georgia Institute of Technology<br>315 Ferst Dr, Rm 3303<br>Atlanta, GA 30332<br> | Monica Garcia-Alloza<br>Division of Physiology, School of Medicine.<br>Universidad de Cadiz. Instituto de Investigacion Biomedica de Cadiz (INIBICA).<br>Cadiz, Spain.<br> |
|---|---|

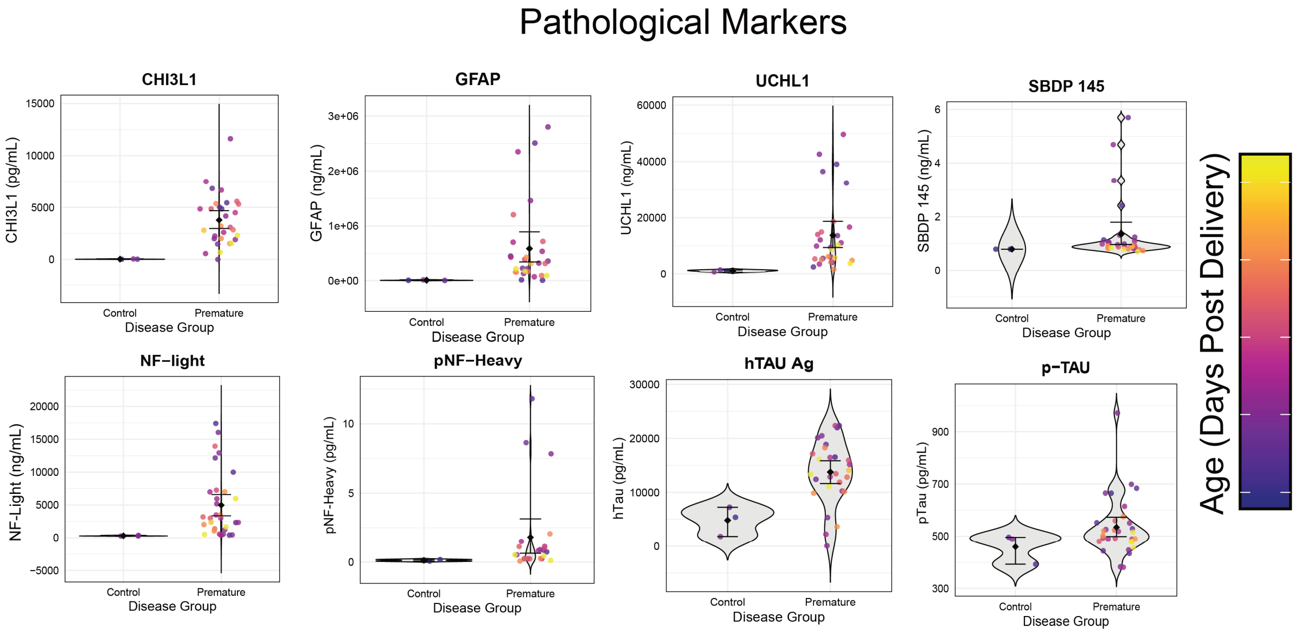


**Figure S1: Neurodegeneration markers increase in premature neonate compared to control (N=4-6 per group, mean ± SEM).** The color bar represents age of the sample ranging from 10 (purple) to 60 (yellow) days post delivery.

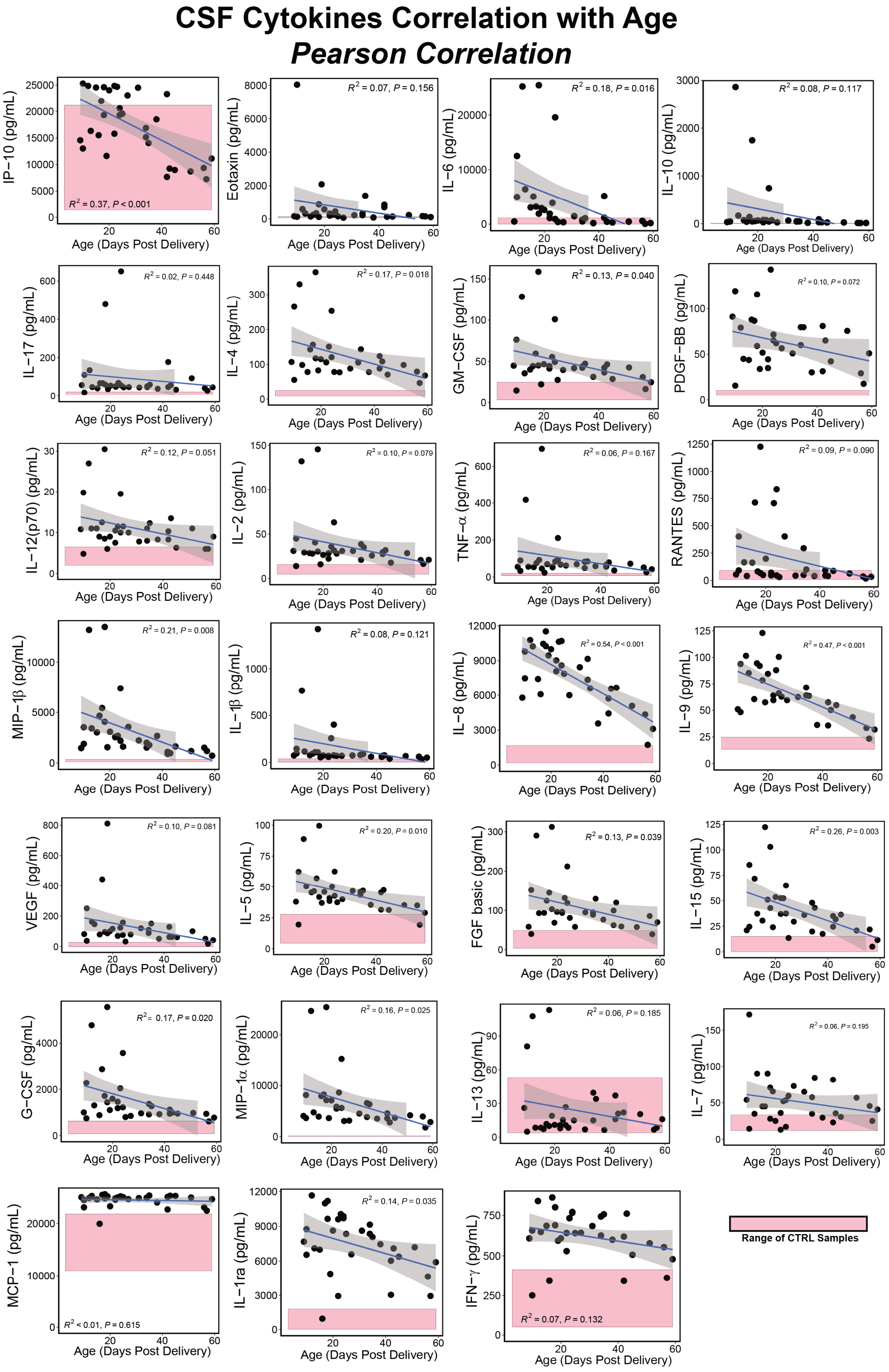


**Figure S2: Longitudinal analysis of CSF cytokine levels.** Pink boxes show range of control patients, grey indicates 95% CI from linear regression, p-value from Pearson correlation.

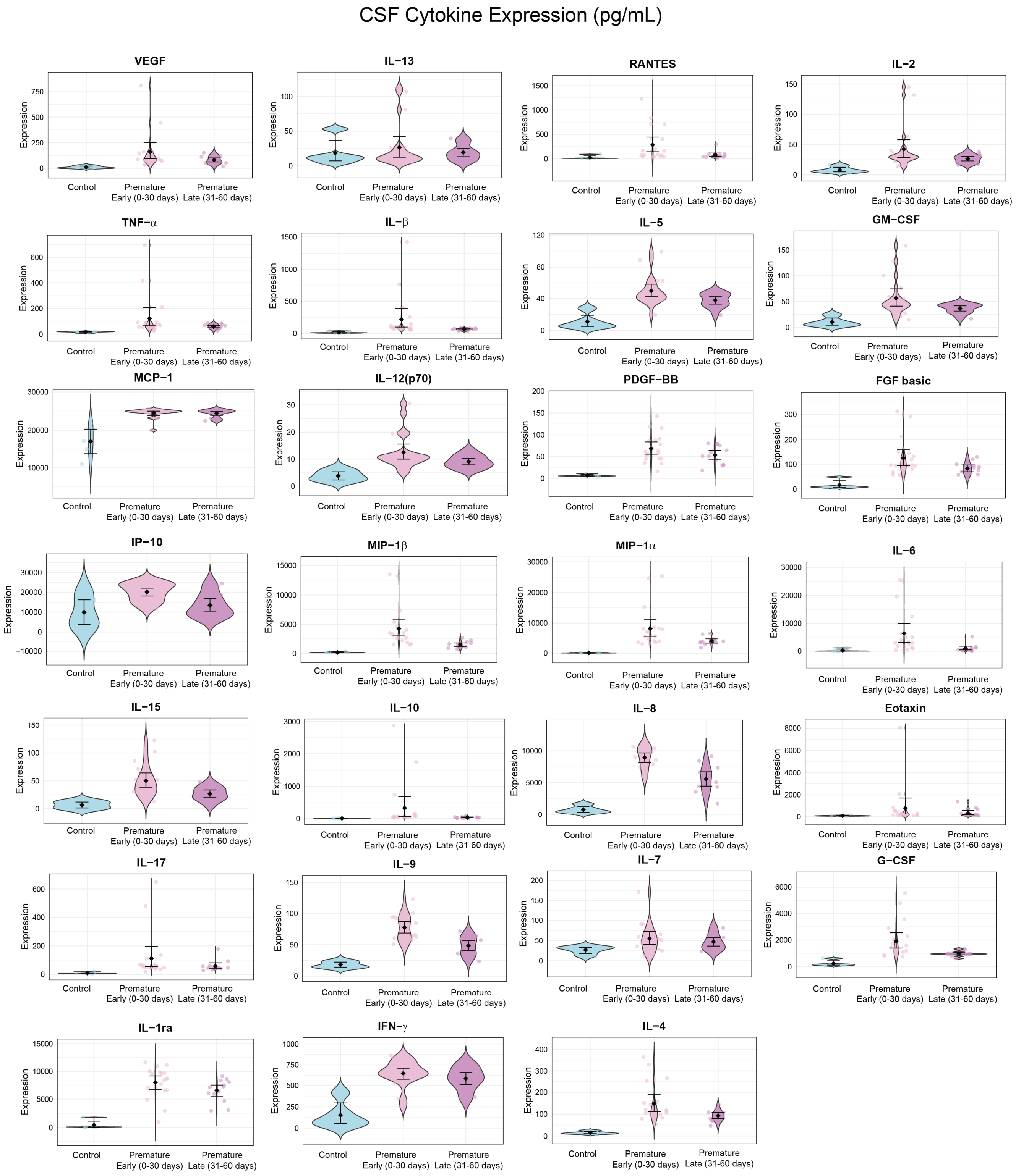


**Figure S3: CSF cytokine levels shown in control, premature (0-30 days) and premature (31-60 days)**. N=4-6 per group, mean ± SEM

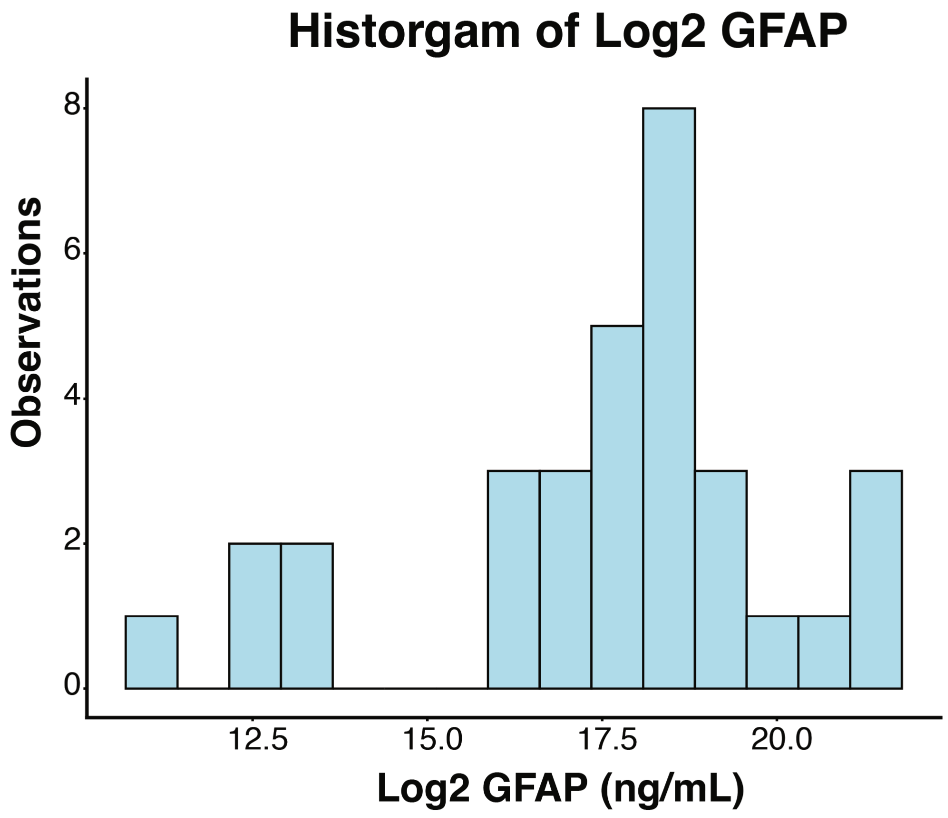


**Figure S4: GFAP expression level is normally distributed after log2 transformation.**
